# Dominant negative OTULIN Related Autoinflammatory Syndrome

**DOI:** 10.1101/2023.03.24.23287549

**Authors:** Sophia Davidson, Yuri Shibata, Sophie Collard, Pawat Laohamonthonku, Klara Kong, June Sun, CIRCA, AADRY, Margaret W.Y. Li, Carolyn Russell, Anna van Beek, Edwin P. Kirk, Rebecca Walsh, Paul E. Gray, David Komander, Seth L. Masters

**Affiliations:** Inflammation Division, The Walter and Eliza Hall Institute of Medical Research, 1G Royal Parade, Parkville, VIC 3052, Australia; Department of Medical Biology, The University of Melbourne, Parkville, VIC 3010, Australia; Ubiquitin Signaling Division, The Walter and Eliza Hall Institute of Medical Research, 1G Royal Parade, Parkville, VIC 3052, Australia; School of Medicine, University of New South Wales, Sydney, Australia; Department of immunology and infectious diseases, Sydney children’s hospital,Randwick, Sydney, Australia; Department of Paediatric Surgery, Sydney Children’s Hospital,Randwick, Sydney, Australia; Department of General Paediatrics, Sydney Children’s Hospital, Randwick, Sydney, Australia; Department of Medical Genetics, Sydney Children’s Hospital,Randwick, Sydney, Australia; NSW Health Pathology Genetic Laboratory, Randwick, Sydney, Australia; University of Western Sydney, Australia

## Abstract

Biallelic loss of function mutations in the linear chain specific deubiquitinase (DUB) OTULIN (OTU Deubiquitinase With Linear Linkage Specificity) result in OTULIN Related Autoinflammatory Syndrome (ORAS). To date all reported ORAS patients have had homozygous or compound heterozygous loss of function mutations, however we identified a patient with a monoallelic heterozygous mutation p.Cys129Ser. Consistent with the ORAS phenotype, we observed accumulation of linear ubiquitin chains, increased sensitivity to TNF induced cell death and dysregulation of inflammatory signalling in both patient cells and in vitro exogenous expression models. Levels of the mutant OTULIN protein were consistent with wild type OTULIN in patient cells and exogenous expression systems and maintained binding capacity to both LUBAC and linear ubiquitin chains. However, even in a heterozygous context this mutant DUB promoted the global accumulation of linear ubiquitin chains. Furthermore, it allowed accumulation of ubiquitin on the linear ubiquitin chain assembly complex (LUBAC). Altered ubiquitination of LUBAC leads to a dysregulation of NF-κB signalling and promotion of TNF induced cell death. By reporting the first dominant negative mutation driving ORAS this study expands our clinical understanding of Otulin mediated pathology.

**Summary:** The dominant negative mutation *OTULIN*:p.Cys129Ser is sufficient to drive ORAS. Mutant OTULIN outcompetes wildtype for interaction with SHARPIN, this increases M1-ubiquitination of LUBAC and other targets, thereby promoting cell death and consequent inflammatory signalling.

## Introduction

Ubiquitination is the reversible post-translational covalent modification in which ubiquitin (Ub) is conjugated to target proteins to modulate their function. Ubiquitin itself can be ubiquitinated to generate poly ubiquitin chains by attachment of the incoming ubiquitin to any of seven different lysine residues (K6, K11, K27, K29, K33, K48, K63) or in a linear manner using the N-terminal methionine (M1). Linear or Met1-linked Ub chains are particularly important for innate immune receptor signalling and control pro-inflammatory signalling through nuclear factor-κB (NF-κB) or mitogen-activated protein kinases (MAPK), and for the inhibition of cell death (Swatek and Komander, 2016; Jahan *et al*., 2021).

Ubiquitin modification is catalysed by the repetitive function of three enzymes: E1 (ubiquitin activating enzyme), E2 (ubiquitin conjugating enzyme), and E3 (ubiquitin ligase). The linear ubiquitin chain assembly complex (LUBAC) is the only E3 enzyme complex identified to date that generates Met1-linked ubiquitin chains (Kirisako *et al*., 2006). LUBAC is composed of three subunits: a large isoform of heme-oxidized iron regulatory protein 2 (IRP2) ubiquitin ligase 1 (HOIL-1L), HOIL-1L interacting protein (HOIP), and SHANK-associated RH domain-interacting protein (SHARPIN) (Kirisako *et al*., 2006; Gerlach *et al*., 2011; Ikeda *et al*., 2011; Tokunaga *et al*., 2011). LUBAC activity is best described in the context of tumour necrosis factor (TNF) signalling, in which LUBAC is recruited to the TNF receptor signalling complex to modify receptor interacting protein kinase 1 (RIPK1) and NF-κB essential modulator (NEMO) with Met1-Ub chains (Haas *et al*., 2009). This promotes NF-κB signalling and inhibits cell death by blocking complex II formation. Mutations in HOIP and HOIL1 in humans and SHARPIN in mice lead to loss of M1 ubiquitin signalling, autoinflammation and immunodeficiency (Gerlach *et al*., 2011; Ikeda *et al*., 2011; Tokunaga *et al*., 2011; Boisson *et al*., 2012; Boisson *et al*., 2015; Oda *et al*., 2019)

Ubiquitination is counteracted by deubiquitinating enzymes (DUBs). In humans, a single DUB selectively and effectively disassembles Met1-linked Ub chains: OTU deubiquitinase with linear linkage specificity (OTULIN) (Keusekotten *et al*., 2013; Rivkin *et al*., 2013). A second DUB, the ubiquitin-specific protease CYLD, comprises a M1-selective catalytic domain (Komander *et al*., 2009; Sato *et al*., 2015); however, full-length CYLD preferentially cleaves K63 chains (Elliott *et al*., 2021). Both OTULIN and CYLD bind HOIP in the LUBAC complex, either directly (OTULIN), or indirectly via SPATA2 (CYLD) (Keusekotten *et al*., 2013; Rivkin *et al*., 2013; Elliott *et al*., 2014; Schaeffer *et al*., 2014; Elliott *et al*., 2016; Kupka *et al*., 2016; Schlicher *et al*., 2016). One role of OTULIN is to remove linear chains from components of the LUBAC complex generated by auto-ubiquitination; however OTULIN within and outside LUBAC (Stangl *et al*., 2019) serves to globally downregulate inflammatory Met1-Ub chains. Consistently, downregulation of OTULIN results in enhanced Met1-linked ubiquitination of LUBAC and its substrates (Fiil *et al*., 2013; Hrdinka *et al*., 2016). Accumulation of linear ubiquitin chains triggers inflammatory signalling (Damgaard *et al*., 2016; Damgaard *et al*., 2019; Damgaard *et al*., 2020) and inhibits LUBAC’s ability to regulate NF-κB signalling and protect against cell death (Fuseya and Iwai, 2021). Interestingly, while loss of OTULIN in immune cells triggers aberrant inflammation and cytokine release (Damgaard *et al*., 2016), some cell types and organs compensate for the loss of OTULIN by downregulating LUBAC; these cells are unable to appropriately respond to cytokine signals and undergo apoptosis (Damgaard *et al*., 2019).

Similar to patients with loss of function mutations in LUBAC, patients harbouring biallelic loss-of-function mutations in *OTULIN* develop a severe auto-inflammatory disease with skin involvement, termed ORAS (OTULIN-related auto-inflammatory syndrome, also known as Otulipenia, OMIM # 617099) (Damgaard *et al*., 2016; Zhou *et al*., 2016; Tao *et al*., 2021; Zinngrebe *et al*., 2022). Furthermore, mice deficient for OTULIN or expressing a catalytically inactive OTULIN mutant (C129A) die mid-gestation due to angiogenic deficits and aberrant cell death mediated by TNFR1 signalling and receptor-interacting protein kinase 1 (RIPK1) kinase activity (Rivkin *et al*., 2013; Damgaard *et al*., 2016). To date all reported ORAS-associated variants have been biallelic loss of function, while heterozygous loss of function variants can compromise patient immune response to certain pathogens such as *Staphylococcus aureus* (*S. aureus*) with the potential to result in severe necrotising disease (Spaan *et al*., 2022). Herein we describe the first heterozygous, dominant negative variant in OTULIN to cause ORAS. This unexpected result demonstrates the importance of understanding the functional consequences of specific mutations, expanding our understanding of mutations causing ORAS and OTULIN biology.

## Results and Discussion

### Patient Phenotype

The patient is a 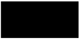 male, born to non-consanguineous 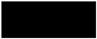 parents. Born at weeks of gestation by emergency caesarean section for maternal eclampsia and required mechanical ventilation for 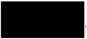after birth. At 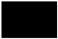 days old he developed peri-umbilical erythema and was diagnosed with a presumed abscess, which was refractory to broad spectrum antibiotics. Surgery was performed and significant inflammation was observed around a urachal remnant, with high numbers of neutrophils seen on histopathological examination, yet no bacteria was cultured from the site. The surgical closure dehisced and he was left with an abdominal wound that failed to close (Figure 1a). Across the period of healing of 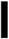 months, the only positive culture was a light growth of methicillin sensitive *S. aureus*.

**Figure 1.**
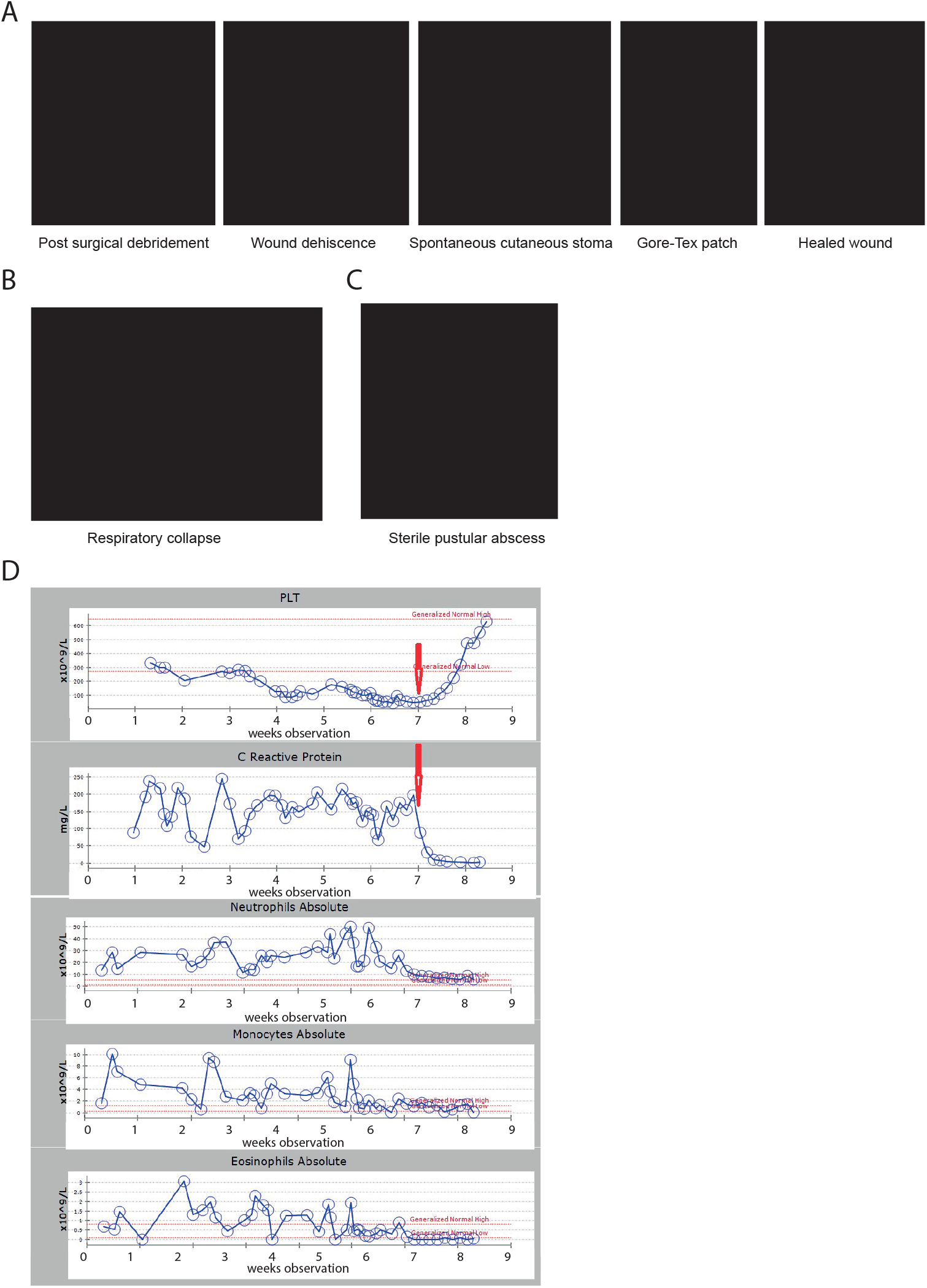
Patient with heterozygous C129S variant in OTULIN exhibits features consistent with ORAS. **(A)** Photographs detailing surgical wound progression. **(B)** Acute respiratory deterioration. **(C)** Sterile pustular abscesses at sites of previous trauma, e.g.: injection site on ankle. **(D)** Frequencies of platelets (PLT), neutrophils, monocytes and eosinophils and concentration of C Reactive Protein (CRP) in patient blood measured weekly. Red dotted lines indicate normal range and red arrow indicates commencement of adalimumab treatment. **(A-C)** Please contact the corresponding author to request access to data.

At 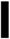 weeks of age, the patient suffered an acute respiratory deterioration (Figure 1b) associated with extensive body edema, hypotension and presumed pulmonary edema. High ventilation pressures were required to maintain lung oxygenation. Again, no organisms were isolated other than a rhinovirus in nasopharyngeal aspirate. Interestingly, corticosteroids were initiated as a supportive measure and the lungs immediately improved. Furthermore, the initiation of corticosteroid treatment correlated with the abdominal wound beginning to heal. This has continued, although he developed a spontaneous cutaneous stoma from his duodenum (Figure 1a). In addition to steroids he was treated with TNF-blockade (Adalimumab) which seemed to improve the rate of healing allowing for a reduced steroid dose, and JAK inhibition (Ruxolitinib). Furthermore, the rate of healing was noted to be remarkably better when he was covered with broad spectrum antibiotic prophylaxis, initially with timentin, and subsequently intravenous and then nasogastric co-trimoxazole. During this time the patient also developed a systolic murmur associated with a 1.2×1.6mm echogenic focus on the tricuspid valve, and was treated for infective endocarditis, but again with no causative organism identified on repeated culture.

Throughout his intensive care unit admission of many months, he developed repeated areas of swelling and erythema consistent with panniculitis, and additionally developed lesions that were believed to represent pathergy, with sterile pustular abscesses forming at the sight of prior trauma, including injection and cannula sites at the ankle and thigh (Figure 1c). Consistent with the abdominal wound healing, these lesions were responsive to corticosteroids.

Investigations demonstrated raised inflammatory markers (CRP 100-200mg/L) and elevated neutrophils (30-50×10^9^/L), monocytes (2-10×10^9^/L), eosinophilia (1-3×10^9^/L) and a persistent thrombocytopaenia (below 100×10^9^/L), which continued until inflammation was controlled with corticosteroids (Figure 1d). The patient also demonstrated an initial hypogammaglobulinemia (IgG =0.59g/L, IgA < 0.07g/L), although this may have related to prematurity. In contrast to the elevated myeloid populations, lymphocyte subsets were largely normal (data not shown). With time he has begun to develop antibodies normally (IgA = 0.31g/L, IgM = 0.61g/L) with IgG still replaced. Biopsy of perilesional skin from the abdominal wound showed heavy acute inflammatory exudate with diffuse suppuration and areas of necrosis. Persistent inflammation and evidence of cell death were considered consistent with an autoinflammatory disease caused by an inborn genetic error.

### Genetic studies

Trio exome sequencing identified a *de novo* variant in OTULIN, ENST00000284274.5:c.386G>C, resulting in a missense change from cysteine to serine at position 129 (p.Cys129Ser). This variant was previously unreported and absent from databases of normal variation, including gnomAD (v2.1.1 and v3.1.2). In silico pathogenicity prediction tools were equivocal with 11 out of 22 queried by the Varsome website consistent with a deleterious effect, a Combined Annotation Dependent Depletion (CADD) score of 24.6, and a moderate physiochemical change (Grantham score = 112). Cysteine 129 is part of the OTULIN catalytic domain and is essential for OTULIN enzymatic activity (Keusekotten *et al*., 2013; Rivkin *et al*., 2013). The amino acid change from cysteine to serine renders the catalytic site inactive, leading to a loss of OTULIN activity. Indeed, a mouse model of a similar variant, C129A OTULIN has been shown to drive an autoinflammatory disease phenotype characterised by aberrant cell death and dysregulation of TNF and the type I interferon (IFNαβ) signalling pathways (Heger *et al*., 2018). We hypothesised that the C129S variant likewise drives autoinflammatory disease. However, as all reported ORAS patient variants result in almost complete loss of OTULIN function, this heterozygous C129S variant may act in a dominant negative fashion to drive disease.

### Heterozygous C129S mutation of OTULIN does not alter protein stability but affects deubiquitinase activity

The OTULIN C129S mutant showed similar protein stability with similar melting temperature to wild type (WT) OTULIN in a Tycho NT.6 assay (Figure 2a), and as expected, OTULIN C129S or C129A variants are devoid of catalytic activity (Supp Fig. 1a). We did not observe a consistent difference in OTULIN transcription or protein expression in patient fibroblasts (Figure 2b,c, Supp Fig. 1b). OTULIN deficiency in certain cell types of ORAS patients can lead to loss of LUBAC components (Damgaard *et al*., 2016), however we did not observe a consistent decrease in any LUBAC component in patient fibroblasts at either the transcript or protein level (Figure 2b,c, Supp Fig. 1b).

**Figure 2.**
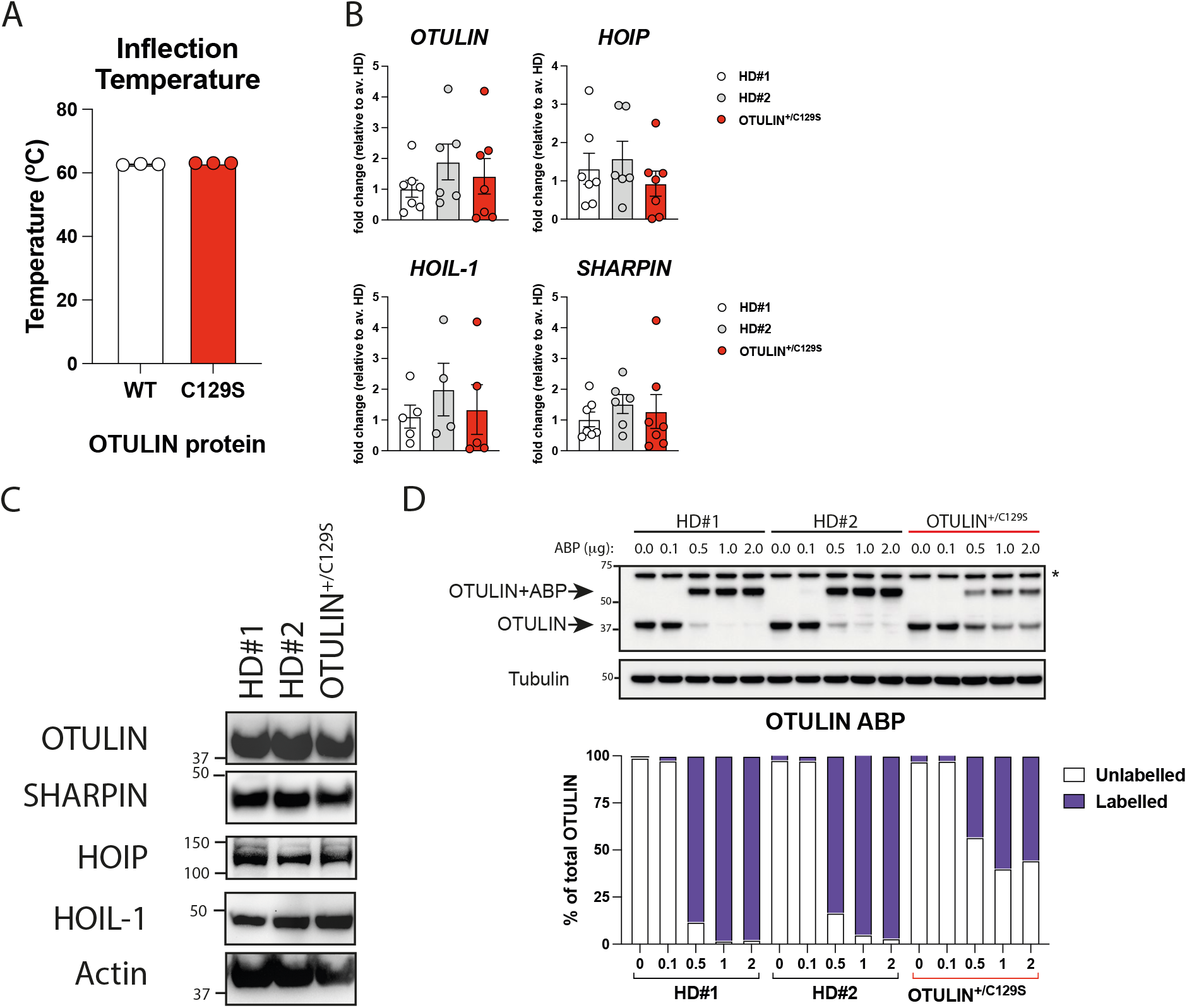
C129S OTULIN mutant protein associates with increased Met1 ubiquitination and cell death in patient cells. **(A)** C129S OTULIN protein thermostability is comparable to WT OTULIN protein, as assessed by Tycho. **(B)** Transcription of *OTULIN, SHARPIN, HOIL-1*, and *HOIP* in HD#1, HD#2 and patient fibroblasts was assessed by qPCR. **(C)** Protein levels of OTULIN, SHARPIN, HOIL-1, HOIP and actin were assessed in healthy donor (HD#1, HD#2) and patient fibroblasts by immunoblot. **(D)** Fibroblast lysates from HD#1, HD#2 and patient lines were treated with the OTULIN ABP at stated concentrations to assess OTULIN activity. OTULIN and Tubulin (loading control) were assessed by immunoblot, arrows indicate unlabelled (OTULIN) and labelled (OTULIN+ABP) OTULIN. * indicates non specific band. Densitometry analysis of labelled and unlabelled OTULIN bands was performed and graphed as a percentage of total signal. **(A**,**B)** Data are pooled from at least three experiments, and statistical significance was assessed by unpaired t tests. **(C**,**D)** Data representative of at least 3 experiments, means +/- SEM.

Whist the C129S mutation did not alter OTULIN protein stability this mutation did affect OTULIN enzymatic activity. A chemically enhanced, linear di-ubiquitin activity based probe (ABP) used to covalently and specifically modify OTULIN in cell lysates (Weber *et al*., 2017) confirmed that in patient fibroblasts, both active and inactive forms of OTULIN are co-expressed, in seemingly equal amounts (Figure 2d).

### C129S mutant OTULIN drives the accumulation of linear ubiquitin chains in the presence of wild type OTULIN

While we did not observe changes in OTULIN or LUBAC protein levels in patient cells we did observe accumulation of linear ubiquitin chains as assessed by GST-NEMO Linear Tandem Ubiquitin Binding Entity (TUBE) pull down (Figure 3a). Linear ubiquitin chain levels were notably higher in patient fibroblasts compared to what was observed healthy donor samples (Figure 3a). Moreover, similar results were observed when we transiently transfected HEK 293T cells with WT or C129S OTULIN (Figure 3b). Importantly, as we performed this experiment in wild type HEK 293T cells, mutant OTULIN drove accumulation of Met-1 chains despite the presence of endogenous OTULIN. Thus, overexpressed C129S OTULIN has a dominant negative effect over endogenous WT OTULIN, demonstrating that this mutation may cause autoinflammatory disease even in a heterozygous context.

**Figure 3.**
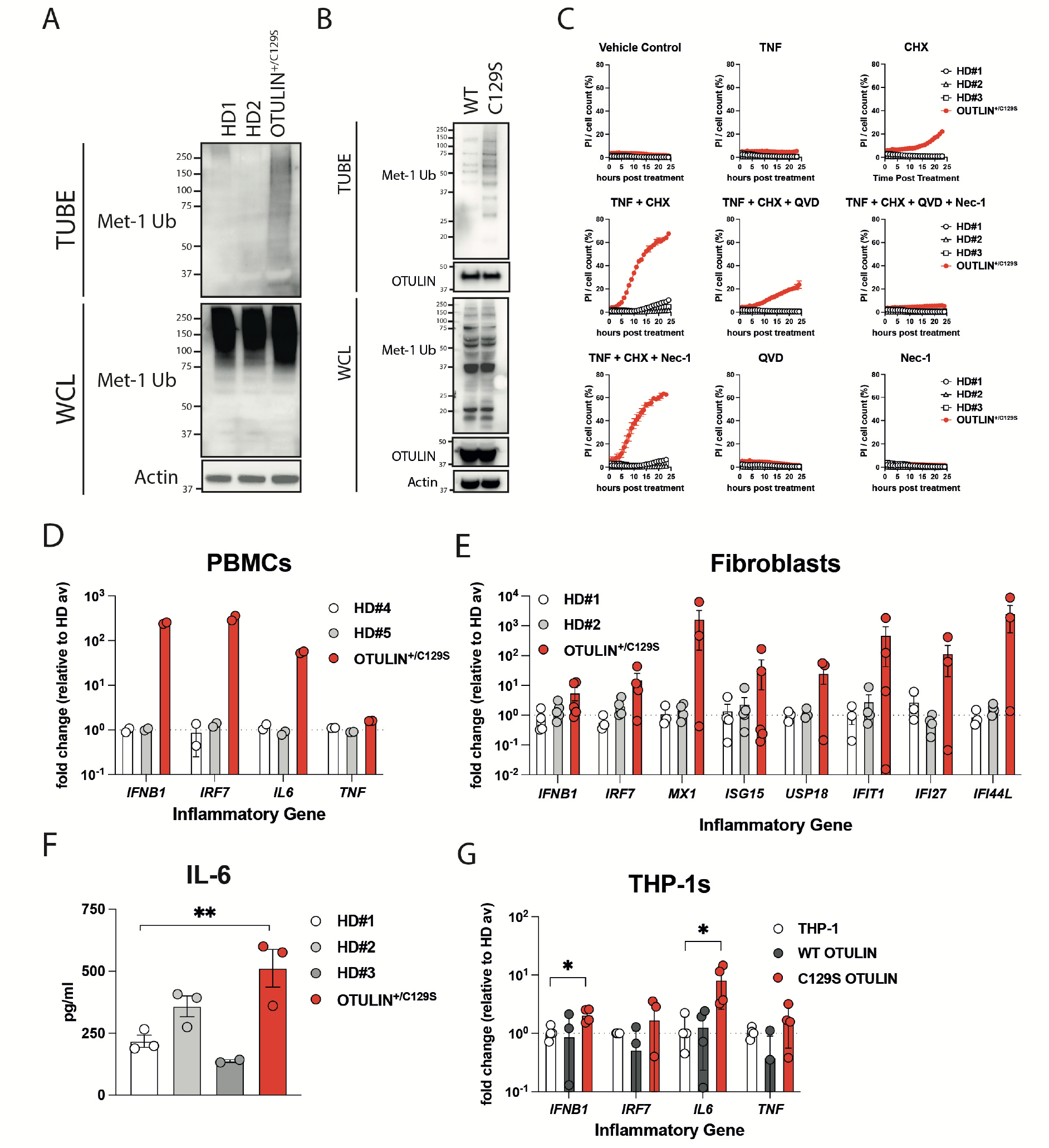
C129S OTULIN mutant protein associates with elevated inflammation. **(A**,**B)** TUBE (tandem ubiquitin binding entity) reagent was used to pull-down polyubiquitin in cell lysates. Linear ubiquitin chains were then specifically probed for in TUBE pull down and whole cell lysate (WCL) samples. Equal loading was confirmed by blotting for actin in the WCL. This was performed in **(A)** lysates from HD#1, HD#2 and patient fibroblasts and in **(B)** lysates from HEK293T cells transiently transfected with either WT or C129S OTULIN. **(C)** HD#1, HD#2 and patient fibroblasts were treated with TNF (100 ng/ml), CHX (50 µg/ml), Q-VD-OPh (10 µM) and Nec-1 (10 µM) or a combination thereof, as indicated. Cell death was measured over a period of 24 hours by cellular inclusion of propidium iodide dye. **(D**,**E**,**G)** Transcription of inflammatory genes including: *IFNB1, IRF7, IL6, TNF, MX1, ISG15, USP18, IFIT1, IFI27* and *IFI44L* was assessed by qPCR in **(D)** patient and healthy donor PBMCs, **(E)** patient and healthy donor fibroblast lines and **(G)** THP-1 cells stably expressing WT or C129S OTULIN or non-transduced controls. Inflammatory genes of interest are expressed as fold change relative to the **(D**,**E)** average of the healthy donors or **(G)** non-transduced THP-1s. **(F)** Secretion of IL-6 from patient and healthy donor fibroblasts were assessed by ELISA. **(A-C)** Data representative of at least 3 experiments. **(D)** Data representative of one experiment, technical replicates graphed. **(E-G)** Data are pooled from at least three experiments, means +/- SEM, and statistical significance was assessed by one way ANOVA, where * indicates P < 0.05.

Collectively, these results are strikingly different to a distinct, homozygous *OTULIN*:pGly281Arg mutant, which destabilises OTULIN and leads to loss of LUBAC components HOIP and SHARPIN in patient fibroblasts (Damgaard *et al*., 2019).

### OTULIN C129S mutation associates with increased sensitivity to TNF induced cell death and spontaneous inflammatory signalling

A recent study by Spaan et al demonstrated that heterozygous *OTULIN* loss of function variants confer a predisposition to sporadic severe necrosis of the skin and lungs, typically, but not exclusively, after infection with *S. aureus* (Spaan *et al*., 2022). Our patient’s phenotype was more pervasive but his wound has been exacerbated by microbial infection. Along with corticosteroid treatment, antibiotics were essential to wound recovery, consistent with bacterial invasion contributing to his phenotype. Given that this patient has a heterozygous variant, we wanted to determine if the patient is suffering from ORAS or OTULIN haploinsufficiency. One of the key cellular differences between OTULIN haploinsufficiency and ORAS, is sensitivity to TNF induced cell death. OTULIN haploinsufficient fibroblasts are not sensitive to TNF induced cell death while fibroblasts from ORAS patients exhibit enhanced cell death via increased complex II formation (Damgaard *et al*., 2019; Spaan *et al*., 2022).

We investigated viability of OTULIN C129S patient fibroblast compared to healthy donor fibroblasts upon treatment with TNF in combination with the protein synthesis inhibitor cycloheximide (CHX). C129S OTULIN patient fibroblasts died at a significantly higher rate compared to healthy donor cells, as assessed by cellular inclusion of propidium iodide dye (Figure 3c). Treatment with the pan caspase inhibitor, QVD significantly reduced TNF-CHX induced cell death, while the RIPK1 kinase inhibitor, necrostatin-1 (Nec-1) only slightly inhibited cell death. Combination treatment of QVD and Nec-1 entirely blocked TNF-CHX induced cell death of patient fibroblasts (Figure 3c). Thus, OTULIN C129S patient fibroblasts die primarily by apoptosis and, to a lesser extent, by necroptosis in response to TNF+CHX stimulation. This matches previous observations made in fibroblasts derived from ORAS patients, despite our patient having different LUBAC/OTULIN levels (as mentioned above), and does not match observations made in OTULIN haploinsufficient individuals (Damgaard *et al*., 2019; Spaan *et al*., 2022).

In contrast to ORAS, patients with heterozygous loss of function mutations in OTULIN do not exhibit spontaneous autoinflammatory disease, and investigation of specific cell types did not reveal any subclinical inflammatory disturbances in these individuals (Spaan *et al*., 2022). Despite our patient having a heterozygous OTULIN variant, we observed a marked increase in transcription of the proinflammatory cytokines, *IFNB1* and *IL6* and to a lesser extent, *TNF* in our patient’s peripheral blood mononuclear cells (PBMCs) (Figure 3d). Along with elevated transcription of *IFNB1*, we also observed increased basal secretion of IL-6 from OTULIN C129S patient fibroblasts compared to healthy donor lines (Figure 3e,f).

*IFNB1* is a member of the type I IFN family, this master cytokine family triggers transcription of hundreds of genes collectively known as Interferon stimulated genes (ISGs). Consistent with elevated *IFNB1* in patient cells, we also observed elevated expression of the ISGs *IRF7, MX1, ISG15, USP18, IFIT1, IFI27* and *IFI44l* in patient fibroblasts, compared to two healthy donors (Figure 3e). Similarly, transcription of *IRF7* was elevated in patient PBMCs compared to healthy donors. However due to material limitations we could not analyse a full panel of ISGs (Figure 3d). Again, this elevated inflammatory gene transcription is consistent with the ORAS phenotype, in which Tao et al recently reported an elevated type I IFN gene signature in the blood of ORAS patients, with loss of OTULIN resulting in accumulation of Ub chains on proteasomal subunits inhibiting proteasome function (Tao *et al*., 2021). In contrast, expression of ISGs in PBMCs from OTULIN haploinsufficient patients were not elevated (Spaan *et al*., 2022). Finally, a similar inflammatory gene signature was observed in THP-1 cells stably expressing the C129S OTULIN mutation compared to THP-1 cells expressing WT OTULIN and the parental cell line (Figure 3g).

Collectively this data demonstrates that the OTULIN C129S variant, although heterozygous, drives the autoinflammatory disease, ORAS. To this point, treatment with a TNF blocking therapy, adalimumab has allowed the patient to begin weaning off corticosteroids. Blockade of TNF signalling dramatically reduced CRP levels and promoted a decrease in numbers of circulating myeloid cells (Figure 1d). Furthermore, adalimumab therapy supported abdominal wound healing, likely due to inhibition of TNF induced cell death. The patient is now 2 months post-op following closure of the wound. A Gore-Tex patch was used to recreate his abdominal wall (Figure 1a) and is currently re-established on full feeds.

It is important to note that this patient continues to exhibit increased susceptibility to bacterial infection. For this reason, he is maintained on co-trimoxazole as an antimicrobial prophylaxis. Discontinuation of antibiotics at various points during the patient’s treatment correlated with a deterioration of abdominal wall healing while re-initiation of antibiotics greatly aided healing. These observations are consistent with there being a microbial contribution to patient pathology, however despite repeated testing, we did not consistently identify a causative pathogen. Decreased or complete loss of OTULIN causes the dysregulation of ubiquitination of caveolin-1, a scaffolding protein which promotes cell surface retention of *S. aureus*’s α-toxin receptor, A Disintegrin and Metalloproteinase 10 (ADAM10). Greater cell surface expression of ADAM10 enhanced α-toxin cytotoxicity leading to cell death (Spaan *et al*., 2022). To date, severe necrotising disease has only been reported in OTULIN haploinsufficient patients, however Spaan et al demonstrated that this dysregulation of caveolin-1 and ADAM10 was common to patients with both heterozygous and homozygous loss of function variants in *OTULIN* (Spaan *et al*., 2022). The increased sensitivity to TNF induced cell death and increased inflammatory gene transcription in OTULIN C129S patient cells, as well as the transfer of these features into OTULIN sufficient cell lines by over expression of the OTULIN C129S mutant (Figure 3c) demonstrate that we are reporting the first case of ORAS caused by a heterozygous mutation. Moreover, study of this patient and his necrotising wound demonstrate that microbes may enhance ORAS disease pathology and addition of antibiotics to the treatment regime may prove beneficial, particularly in the context of treatment with therapeutics which block the key antimicrobial cytokine, TNF.

To counter inflammatory signalling driven by elevated type I IFNs observed in patient cells the JAK inhibitor ruxolitinib was also trialled in this patient. However, the discontinuous bowel wound may impede absorption and so the exact benefit of this drug is currently unknown. Given that ORAS patients have been demonstrated to have an ISG signature and the high success of JAK inhibitor treatment in other autoinflammatory diseases which have an ISG signature (Sanchez *et al*., 2018) it would be of interest to determine if ruxolitinib therapy could support complete cessation of steroids.

### C129S OTULIN mutation allows for accumulation of Met-1 ubiquitin chains on LUBAC

The data above establish that a heterozygous C129S mutation in OTULIN is drives an ORAS phenotype. In contrast to other ORAS variants which lead to loss or substantial decrease in OTULIN protein levels, C129S OTULIN has comparable stability to WT OTULIN and total OTULIN protein levels in patient fibroblasts are similar to the levels observed in healthy donors (Figure 2a,c). We observed similar levels of OTULIN pulled down in the TUBE assay in HEK cells transiently over-expressing both WT and C129S OTULIN (Figure 3b), implying that C129S OTULIN can bind to linear ubiquitin chains; yet as Supp Figure 1a shows, C129S OTULIN cannot cleave linear ubiquitin chains. Thus, the C129S mutant OTULIN is likely to be acting in a dominant negative manner to inhibit WT OTULIN function and driving inflammatory disease.

We hypothesised that OTULIN C129S was binding to Met-1 Ub chains and protecting these chains from hydrolysis by wild type OTULIN. OTULIN C129A is the highest affinity Met1-diubiquitin receptor described to date with a binding constant of 112 nM (Keusekotten *et al*., 2013) and OTULIN C129S likely similarly binds, yet cannot cleave Met1-linked Ub chains. However, even at a 5:1 molar ratio of OTULIN C129S to WT, all di-Ub was cleaved in vitro, likely reflecting the dynamic nature of the interaction (Figure 4a). Interestingly, these in vitro results are in contrast with conditions in patient cells in which Met1-linked chains do accumulate in the heterozygous 1:1 setting.

**Figure 4.**
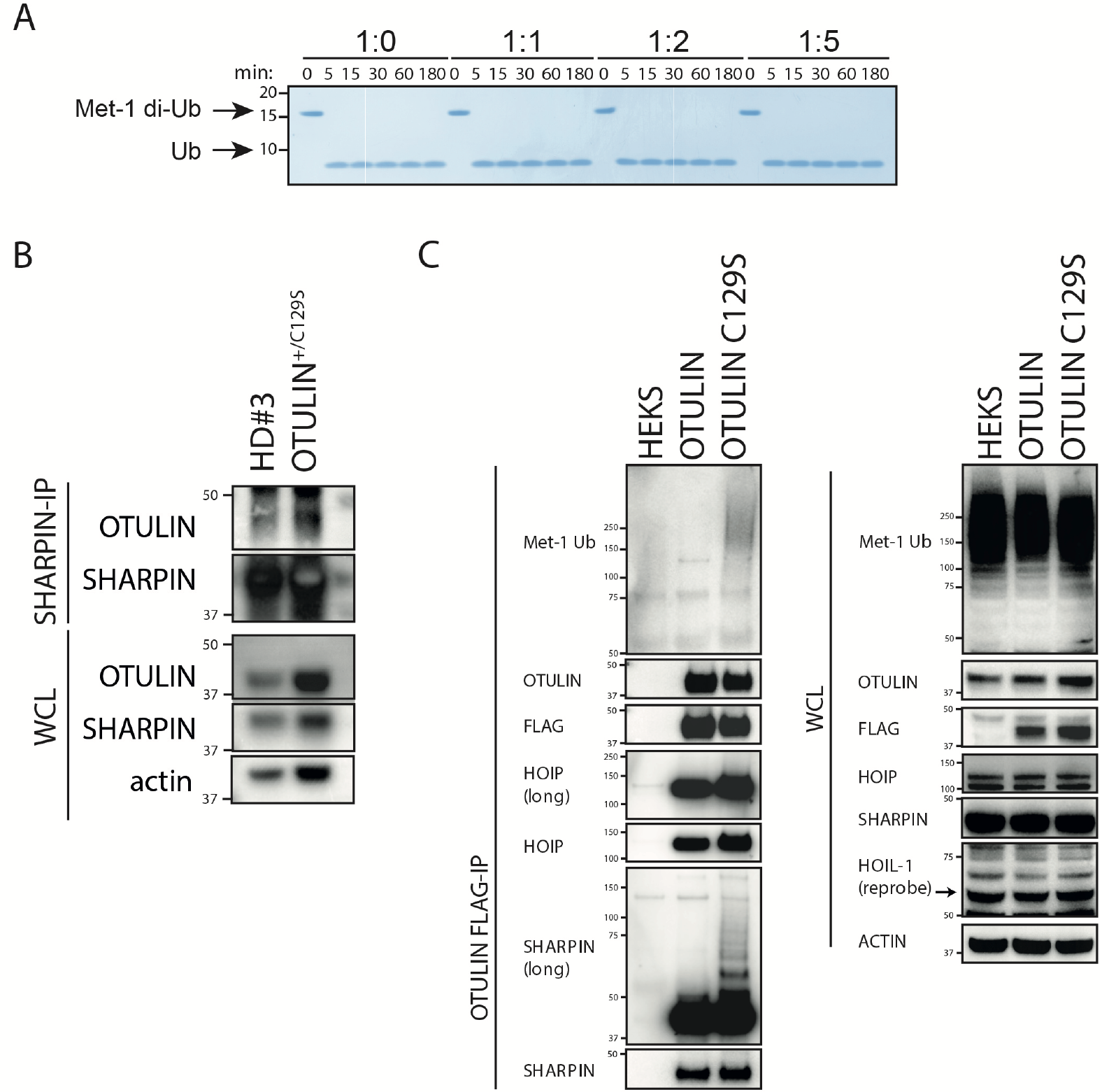
C129S variant renders OTULIN catalytically inactive. **(A)** Cleavage of Met1-linked di-Ub was assessed in competitive settings. Increasing concentrations of C129S OTULIN were incubated with WT OTULIN Met1-linked di-Ub and for 0 to 180 minutes. **(B)** SHARPIN-IP and WCL lysates from HD#3 and patient fibroblasts were immunoblotted for OTULIN, SHAPRIN, and actin (loading control). **(C)** FLAG tagged WT and C129S OTULIN were stably expressed in HEK293T cells. FLAG-IP and WCL lysates from WT OTULIN, C129S OTULIN and parental HEK293T cells were immunoblotted for Met-1 Ub, OTULIN, FLAG, HOIP, SHARPIN, HOIL-1 and actin (loading control).

To understand where the stabilised ubiquitin chains are located, we investigated some of the known OTULIN substrates. OTULIN binds to LUBAC itself and inhibits autoubiquitination of the complex (Elliott *et al*., 2014; Schaeffer *et al*., 2014); with inactive OTULIN, ubiquitinated LUBAC components are readily observed. To test whether this observation holds true in a heterozygous setting, we tested for increased autoubiquitination of LUBAC components, which may lead to inhibition of LUBAC function (Heger *et al*., 2018).

We assessed OTULIN binding to the LUBAC complex in patient and healthy donor fibroblasts by immunoprecipitation (IP) of SHARPIN and observed increased interaction of OTULIN and LUBAC in patient fibroblasts (Figure 4b). Consistent with these results, IP of FLAG tagged OTULIN C129S and WT stably expressed in HEK cells revealed that SHARPIN and to a lesser extent, HOIP, immunoprecipitated with OTULIN C129S were more heavily ubiquitinated compared to the HOIP and SHARPIN pulled down by WT OTULIN (Figure 4c). It is interesting that in this context, active wild type OTULIN which is also present in HEK cells, is unable to revert SHARPIN polyubiquitination status. Additionally, the overall amount of Met-1 ubiquitin chains immunoprecipitated by OTULIN C129S was elevated compared to OTULIN WT (Figure 4c). Thus, the C129S OTULIN heterozygosity leads to accumulation of Met-1 ubiquitin chains on LUBAC complexes, which may drive ORAS in patients. The accumulation of Met1-linked polyubiquitin on LUBAC and other substrates acts as an inflammation trigger and increases sensitivity to drive TNF induced cell death and uncontrolled autoinflammatory signalling.

## Conclusion

We have demonstrated the mechanism by which a heterozygous variant in OTULIN can act in a dominant negative manner to cause ORAS. Considering the recently published phenotype of OTULIN haploinsufficient patients it is surprising that this patient suffers from ORAS. Our patient’s phenotype is likely due to the unique mutation in the catalytic domain of OTULIN, which changes OTULIN into an inactive, Met1-linkage specific ubiquitin receptor without affecting its stability, cellular levels or the OTULIN/LUBAC interaction. In contrast, individuals with a different variant in the OTULIN catalytic triad, N341D, do not present with ORAS, but have OTULIN haploinsufficiency related complications or no phenotype (Spaan *et al*., 2022). The N341D mutation decreases OTULIN affinity to negatively charged proteins such as Ub (Keusekotten *et al*., 2013). It is likely that this decreased affinity results in a loss of function OTULIN rather than a dominant negative OTULIN, as observed with the C129S mutation.

This is the first report of classical ORAS caused by a heterozygous variant. Importantly, understanding the disease-causing effects of this mutation resulted in patient diagnosis and effective, lifesaving treatment. Our study demonstrates the importance of understanding the functional implications of a variant for clinical diagnosis and expands our understanding of variants causing ORAS and OTULIN biology.

## Materials and Methods

### Cell culture, reagents, and inhibitors

Human THP-1 cells were obtained from the American Type Culture Collection (catalog no. TIB-202) and cultured in RPMI 1640 [prepared in-house, RPMI 1640 powder (Life Technologies), 23.8 mM sodium bicarbonate (NaHCO_3_) (Merck), 1 mM sodium pyruvate (C_3_H_3_NaO_3_) (Sigma-Aldrich), penicillin (100 U/ml; Sigma-Aldrich), and streptomycin (100 μg/ml; Sigma-Aldrich)] supplemented with 10% fetal calf serum (FCS) (Sigma-Aldrich). THP-1s were cultured at 37°C in a humidified atmosphere with 10% CO_2_. Human embryonic kidney (HEK) 293T cells were maintained in Dulbecco’s modified Eagle’s medium (DMEM) (Gibco), 40 mM NaHCO_3_, penicillin (100 U/ml; Sigma-Aldrich), and streptomycin (100 μg/ml; Sigma-Aldrich) supplemented with 10% FCS. HEK293T cells were cultured at 37°C in a humidified atmosphere with 5% CO_2_.

### Patient samples

Patient PBMC samples and fibroblast cell line were generated from a peripheral blood sample and skin biopsy from the patient at 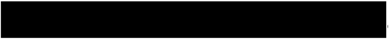after obtaining consent HREC/15/MonH/31 and HREC/15/SCHN/346. Some aspects of patient data have been redacted from this manuscript, if required, please contact the corresponding author to request access to those data. The HD#1 fibroblast line was generated from a skin biopsy at the National Institutes of Health (NIH), NIH Institutional Review Board–approved natural human history study (www.clinicaltrials.gov/ct2/show/NCT02974595) NIH #17-I-0016 “Studies of the natural history, pathogenesis, and outcome of autoinflammatory diseases”. HD#2 and HD#3 fibroblast lines were purchased from the American Type Culture Collection and Lonza respectively. HD#4 and HD#5 PBMC samples were generated from blood draws conducted on adults participating in the Volunteer Blood Donor Registry (VBDR) at WEHI, Human ethics number: HREC ID 18/07. PBMCs were isolated from whole blood using Ficoll (GE Healthcare), frozen in FCS with 10% DMSO, and stored in liquid nitrogen (long term) or -80°C (short term) until use. Fibroblasts were cultured in 10% FCS DMEM at 37°C in a humidified atmosphere with 5% CO_2_. All participants, or their legal representatives, consented to take part in this study and to have the results of this research published.

### Plasmid mutagenesis

We purchased a custom pLV plasmid expressing N terminal FLAG tagged OTULIN and mCherry reporter from Vector Builder. A C129S OTULIN plasmid was obtained via site-directed mutagenesis of the pLV FLAG-OTULIN plasmid using the QuikChange Lightning Kit (Agilent Technologies) using the following mutagenesis primers: FWD: AGTCCGTGGTGATAATTACTCTGCACTGAGGGCC and REV: GGCCCTCAGTGCAGAGTAATTATCACCACGGACT, designed using the Agilent Primer Design Program, synthesised by Integrated DNA technologies. Successful mutagenesis of TGT>TCT was confirmed via Sanger Sequencing.

### Transient Transfection of HEK 293T cells

4×10^6^ HEK 293T cells were seeded into 15cm^2^ dishes (Corning) and cultured overnight. The next morning 12µg of plasmid C129S OTULIN (see Plasmid mutagenesis) or wild type OTULIN expressing plasmids (VectorBuilder) DNA was transfected into HEK 293T cells using FugeneHD (Promega) diluted in OptiMEM (Thermo Fisher Scientific), according to manufacturer’s instructions (3µL of FugeneHD/1µg of DNA). 24hrs later, transfection was removed and replaced with fresh 10% FCS, DMEM. Transfection efficacy was confirmed by assessing mCherry fluorescence using ZOE™ Fluorescent Imager (Biorad). 24hrs after (i.e.: 48 hours post transfection) cells were washed with PBS and lysed for TUBE pull down.

### Stable expression of OTULIN and C129S OTULIN in cell lines

Third generation lentiviral constructs (pLV) for wild type OTULIN (VectorBuilder) and C129S OTULIN (see plasmid mutagenesis) were used to generate lentivirus as described previously (Balka et al., 2020). Breifly, 1.5×10^6^ HEK293T cells were seeded into a 10cm^2^ dish (Corning), after confirmation of cell adherence, HEK293T cells were transiently transfected with pLV OTULIN plasmids, pMDL (packaging), RSV-REV (packaging) and VSVg (envelope) using LipoFectMax (ABP biosciences), diluted in OptiMEM, to generate lentiviral particles. The cell culture supernatant was replaced 24hrs later with appropriate media (i.e.: 10% FCS RPMI for THP-1 cells or 10% FCS DMEM for HEK293T cells). Supernatant was collected a further 24 hours later and filtered through a 0.45µm filter prior to transduction of cell lines. For transduction of THP-1 and HEK293T cells 0.5-3×10^6^ cells were centrifuged with the lentivirus in the presence of polybrene (Sigma-Aldrich) at 839 x g for 3 hours at 32°C and cultured at 37°C overnight. Cells were washed x3 with media to remove all lentivirus. Effectively transduced cells were subsequently selected for by fluorescence activated sorting of mCherry positive cells to generate stable cell lines carrying FLAG tagged OTULIN or FLAG tagged C129S OTULIN.

### Tycho Measurement for Inflection Temperature

The proteins were diluted to 1.0 μM in Tris buffer (20 mM Tris-HCl (pH 7.4), 150 mM NaCl, and 1 mM DTT). Thermal shift assays were performed using Tycho NT.6 (NanoTemper Technologies). The inflection temperatures of each protein were calculated by the tycho NT.6 software.

### Real-time qPCR for cell lines

5×10^6^ PBMCs, 0.5-1×10^6^ THP-1 cells or confluent wells of fibroblasts were lysed using TRIzol reagent (Invitrogen) for 5 minutes at room temperature. Samples were either stored at -80oC or immediately processed. RNA extraction were performed as per the manufacturers’ instructions. cDNA was generated from 0.3 to 1 μg of total RNA using the Invitrogen SuperScript III First-Strand Synthesis System for reverse transcription PCR (RT-PCR) (Invitrogen). qPCR was performed using SYBR Green/ROX qPCR Master Mix (Thermo Fisher Scientific) on a ViiA 7 Real-Time PCR system (Thermo Fisher Scientific). Primers for genes of interest and housekeeping genes were used as described previously (Davidson *et al*., 2022) or listed in Table 1. Each sample was run in duplicate, and samples were normalized using the housekeeping gene *ACTIN*. Results were analysed using the ΔΔCt method and represented as fold change of the average of HD samples from the same experiment.

**Table 1:**
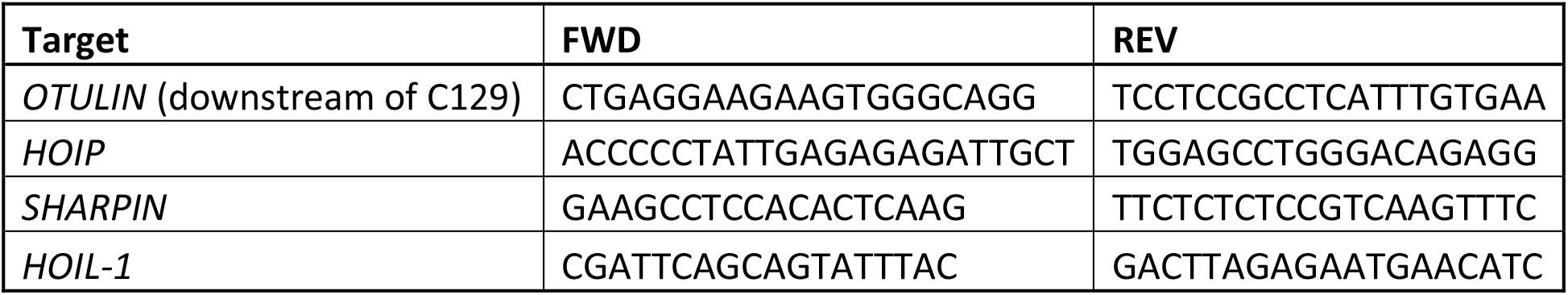
qPCR primer sequences used in this study to assess transcription of *OTULIN* and LUBAC components.

### Immunoblotting

0.3×10e5 fibroblasts were seeded in 6 well plates (corning) and cultured for 18-24 hours prior to lysis. Fibroblasts were lysed in 1× radioimmunoprecipitation assay (RIPA) buffer [1% Triton X-100, 20 mM tris-HCl (pH 7.4), 150 mM NaCl, 1 mM EDTA, 0.5% sodium deoxycholate, 3.5 mM SDS, 10% glycerol, 10 mM NaPPi, 5 mM NaF, and 1 mM Na_3_VO_4_] supplemented with 1 mM phenylmethylsulfonyl fluoride and cOmplete protease inhibitors (Roche Biochemicals) for 30 min at 4°C. Samples were processed through Pierce centrifuge columns (Thermo Fisher Scientific) to remove DNA. After addition of reducing SDS–polyacrylamide gel electrophoresis (SDS-PAGE) sample loading buffer [1.25% SDS, 12.5% glycerol, 62.5 mM tris-HCl (pH 6.8), 0.005% bromophenol blue, and 50 mM dithiothreitol] and denaturation at 95°C for 5 to 10 min, samples were separated on Novex 4 to 12% precast SDS-PAGE gels (Thermo Fisher Scientific) with MES running buffer (Thermo Fisher Scientific) and subsequently transferred onto polyvinylidene difluoride membrane (Millipore). Membranes were blocked in 5% skim milk in tris-buffered saline (TBS) containing 0.1% Tween 20 (Sigma-Aldrich) (TBST) before overnight incubation with specific primary antibodies in 5% bovine serum albumin (BSA) (Sigma-Aldrich) or 5% skim milk in TBST at 4°C: anti-OTULIN [Cell Signaling Technology (CST, #14127), anti-SHARPIN (CST, #4444), anti-HOIP (CST, #99633), HOIL-1 (Sigma Aldrich, clone 2E2), anti–Met-1 (CST, #4560), or anti–actin–horseradish peroxidase (HRP) (1:10,000; Santa Cruz Biotechnology, clone C4, sc-47778). All listed primary antibodies were used at 1:1000 unless otherwise stated. Membranes were washed three times with TBST and incubated with appropriate HRP-conjugated secondary antibodies and washed again three times. Finally, membranes were developed (Chemiluminescent HRP Substrate, Millipore) and imaged using the ChemiDoc Touch Imaging System (Bio-Rad) or film (GE Healthcare or Thermo Fisher Scientific). Band densitometry was assessed using ImageJ2, version 2.9.0/1.53t software.

### Assessment of OTULIN activity using ABP

Fibroblast samples were lysed in lysis buffer (25 mM HEPES pH 7.5, 150 mM NaCl, 0.2% NP-40, 10% glycerol, 1 mM DTT) supplemented with PhosSTOP phosphatase inhibitor (Roche) and without protease inhibitor cocktail. Lysates were divided into aliquots (20-50μL) reaction. 20µg/ml or stated concentrations of OTULIN ABP were added to the samples and incubated at room temperature for 30 min. Reactions were stopped by boiling in reducing sample buffer and analysed by as per described in immunoblotting section. Band densitometry was assessed using ImageJ2, version 2.9.0/1.53t software.

### TUBE pull down

Endogenous polyUb conjugates were assessed in fibroblast lines using TUBE affinity reagents. Briefly, TUBE lysis buffer (20 mM Na_2_HPO_4_, 20 mM NaH_2_PO_4_, 1% NP-40, 2 mM EDTA) was supplemented with 1 mM DTT, cOmplete protease inhibitors (Roche Biochemicals), and 50 μg/ml of GST-TUBE1 (Lifesensors, Malvern, PA). HD and patient fibroblasts were plated in 15cm^2^ dishes at 4×10^6^ cells/dish and cultured for 48 hours. Fibroblasts were lysed in 1ml of ice-cold TUBE lysis buffer and processed as described previously (Hjerpe *et al*., 2009). After isolation, WCL and TUBE pull down samples were boiled in reducing sample buffer and analysed by as per described in immunoblotting section.

### Analysis of cell death

To measure cell death levels, fibroblast cell lines were seeded at 2×104 cells per well in a 48-well flat-bottomed plate. 18-24 hours later, cells were treated with either one single treatment or one combination treatment. Single treatments were 100 ng/ml TNF, 50 μg/ml CHX, 20 μM QVD, 20 μM DMSO and 10 μM Nec-1. Combination treatment were TNF+CHX, TNF+CHX+QVD, TNF+CHX+QVD+Nec-1, and TNF+CHX+Nec-1. PI was added to cell media and cells were imaged using an Incucyte, every hour over 24 hours. Cell death was graphed as PI Object Count / Phase Object Count (%).

### Cytokine measurements

To measure protein levels of IL-6 fibroblast cell lines were seeded at 5×10^3^ cells per well in 96-well flat bottomed plate. 18-24 hours later, culture supernatants were collected, and IL-6 concentration were measured by ELISA using the Human IL-6 Quantikine ELISA Kit, R&D Systems, as per the manufactures’ instructions.

### Generation of recombinant OTULIN proteins

The full-length OTULIN construct (WT and C129A mutant) has been described previously (Keusekotten et al, 2013). OTULIN mutant (C129A and C129S) was generated in pOPINB by site-directed mutagenesis using In-Fusion HD Cloning system with inverse PCR. OTULIN constructs were expressed in Rosetta2 (DE3) pLacI cells. Cells were grown at 37 °C in 2xYT medium with 50 μg/ml kanamycin and 34 μg/ml chroramphenicol to an OD_600_ of 0.6. The cultures were cooled to 18 °C before overnight induction with 200 μM of IPTG. Cells were resuspended in lysis buffer (50 mM Tris-HCl (pH 7.4), 150 mM NaCl, 2 mM β-mercaptoethanol, DNase I, lysozyme, and protease inhibitor cocktail) and lysed by sonication. Lysates were clarified by centrifugation at 19,500 rpm for 30 min, and applied to Cobalt resin for gravity flow purification. The His_6_ tag was cleaved overnight with His-3C protease during dialysis (20 mM Tris-HCl (pH 8.0), 50 mM NaCl, and 2 mM β-mercaptoethanol). Cleaved proteins were reapplied to cobalt resin, and flow-through was collected. The proteins were further purified by anion exchange chromatography (Resource Q, GE Healthcare). Eluted proteins were subjected to size exclusion chromatography (HiLoad 16/600 Superdex 75, GE Healthcare) in SEC buffer (20 mM Tris-HCl (pH 7.4), 150 mM NaCl, 4 mM DTT). Purified proteins were concentrated and flash-frozen prior to storage at -80°C.

### Ubiquitin chain cleavage assay

Qualitative DUB assays were performed as previously described (Keusekotten et al, 2013). Briefly, OTULIN WT and mutants (C129A and C129S) were diluted to a 2x stock concentration in 20 mM Tris-HCl (pH 7.4), 150 mM NaCl, and 10 mM DTT, and incubated with 1 μM of Met1-diUb in the buffer (50 mM Tris-HCl (pH 7.4), 50 mM NaCl, and 5 mM DTT) at 37 °C. The samples were taken at different time points and mixed with 4x LDS sample buffer to stop the reaction. The samples were resolved by SDS-PAGE and visualised by Coomassie staining. For competitive cleavage assays, the indicated concentration of OTULIN (C129A) was pre-incubated with 5 μM of Met1-diUb for 5 min on ice, and 0.5 nM of WT OTULIN was added to the reaction.

### Immunoprecipitation

For Endogenous SHARPIN IP, HD and patient fibroblasts were plated in 15cm^2^ dishes at 4×10^6^ cells/dish and cultured for 48 hours. 5 µg of polyclonal SHARPIN antibody (Proteintech, 14626-1-AP) was bound to 50 µl Dynabeads (Thermo Fisher Scientific) per sample by incubating for 2 hours at 4°C on rotator. Beads were washed x3 with PBS and resuspended in 500 µL PBS. To crosslink antibody to beads, 5 mM of BS3 (Thermo Fisher Scientific) was added and incubated for 30 min at room temperature on a rotator. Crosslink reaction was halted by addition of 1M Tris-HCl and beads were incubated for a further 15 min at room temp on rotator. Beads were washed x3 with DISC buffer (20 mM Tris-HCl, 150 mM NaCl, 2 mM EDTA, 1% Triton X-100 and 10% Glycerol) supplemented with protease inhibitor cocktail tablet (Roche) and Phos-STOP phosphatase inhibitor tablet (Roche). Cells were lysed in DISC lysis buffer for 30 minutes on ice. Insoluble material was removed by centrifugation at 13,000 rpm at 4°C for 30 min. 50 μl of each sample was kept for WCL and processed as per immunoblot samples. SHARPIN antibody conjugated beads was added per 1 ml of lysate and incubated at 4°C overnight on a rotator. Samples were washed 3× with DISC buffer and resuspended in 30 μl of sample buffer. Samples were then boiled for 5 min and subjected to immunoblotting (as per the “Immunoblotting” section).

## Data Availability

All data produced in the present study are available upon reasonable request to the authors

## Author contributions

EPK and RW identified the variant in Otulin. CR, AVB and MWYL were involved in patient treatment, together with PEG. SD, YS, SC, PL, KK and JS were involved in functional validation studies. SD, YS, PEG, DK and SLM wrote the manuscript with input from all authors. PEG, DK and SLM were involved in supervision and conception of the study together with SD and YS.

## Acknowledgments and funding

The authors thank Saskia Vadder (University of Bonn) for technical assistance. This work was supported by grants from the Australian National Health and Medical Research Council (2003159, 2003756; S.L.M.), fellowships from the Victorian Endowment for Science Knowledge and Innovation (S.L.M.), the HHMI-Wellcome International Research Scholarship (S.L.M.) and the Sylvia and Charles Viertel Foundation (S.L.M.). S.D. acknowledges funding from NHMRC grants (GNT1143412 and GNT2003756).

## Figures

**Supp Figure 1.**
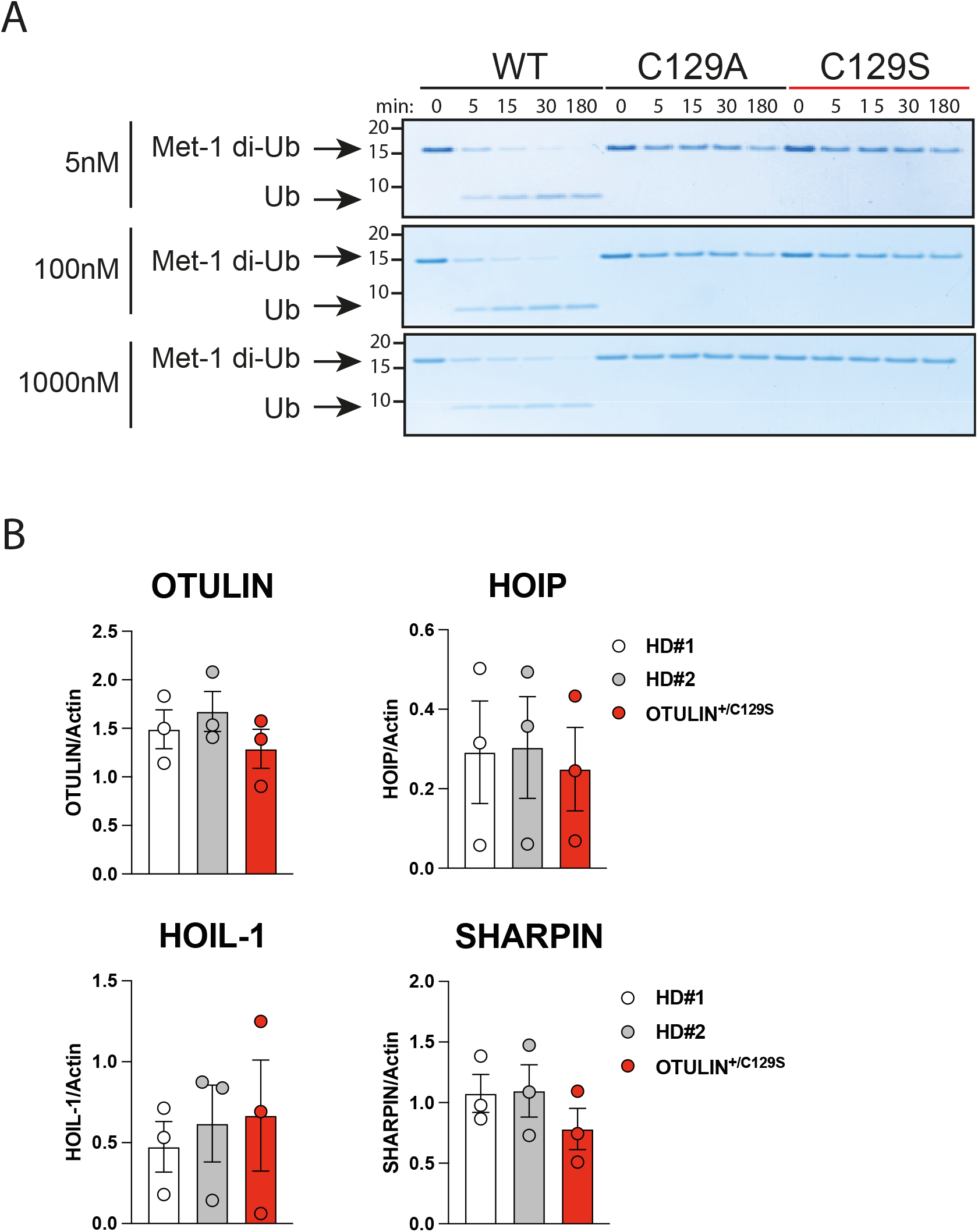
C129S variant renders OTULIN catalytically inactive. **(A)** Recombinant WT OTULIN and C129A and C129S mutant OTULIN proteins were incubated at stated concentrations with Met1-linked di-Ub chains. Cleavage capacity was assessed from 0 to 180 minutes. Arrows indicate uncleaved Met-1 di-Ub and cleaved Ub. **(B)** Fibroblast lysates from HD#1, HD#2 and patient lines assessed for protein levels of OTULIN, HOIP, HOIL-1, SHARPIN and Actin (loading control) were assessed by immunoblot. Densitometry analysis protein expression was performed and graphed as a function of actin loading control. Data are pooled from at least three experiments, means +/- SEM, and statistical significance was assessed by one way ANOVA

